# Urinary peptidomic signatures predict overall and progression-free survival in patients with bladder cancer

**DOI:** 10.1101/2025.10.28.25338944

**Authors:** Melika Ahangar, Harald Mischak, Napoleon Moulavasilis, Konstantinos Stravodimos, Forouzandeh Mahjoubi, Joachim Jankowski, Maria Frantzi, Antonia Vlahou

**Author notes:** **Corresponding Author:** Dr. Maria Frantzi, Mosaiques Diagnostics GmbH, Rotenburger Str. 20, 30659 Hannover, Germany.

## Abstract

Clinicopathologic calculators for bladder cancer moderately predict survival and fail to depict the underlying molecular phenotype. We applied urinary capillary electrophoresis–mass spectrometry (CE–MS) to identify prognostic signatures linked to bladder cancer outcome. In a discovery cohort (n=131; mean follow-up 623 days), 114 survival-associated peptides, predominantly collagen fragments, were significant prognostic factors of survival and were integrated into a classifier (BC110), resulting in an accuracy of 0.80 (p-value < 0.001). Validation of the classifier in an independent cohort (n=102; mean follow-up 1605 days) confirmed correlation with survival (AUC: 0.78; p-value=0.03). Survival analysis using the BC110 scores resulted in significant prediction of both overall (p-value<0.0001) and progression-free survival outcome (p-value < 0.0001). To test biological plausibility, a previously reported collagen-focused model (COL210) was subsequently investigated and demonstrated concordant prognostic separation, reinforcing extracellular matrix remodeling as the underlying signal. These urine-based classifiers enable non-invasive risk stratification and may complement guideline calculators by identifying high-risk patients for adjuvant therapy and low-risk groups for reduced surveillance, potentially lowering reliance on repeated cystoscopy.

**Significance Statement:** Accurate non-invasive risk stratification in bladder cancer remains a major unmet clinical need, given the disease’s high recurrence rates and the need for cystoscopic surveillance. We applied a standardized urinary peptidomics workflow using capillary electrophoresis–mass spectrometry (CE–MS) to develop a machine learning based classifier (BC110) enriched in collagen and other extracellular matrix (ECM) components, in a discovery cohort (n=131) of mainly NMIBC patients. In an independent validation cohort (n=102), BC110 achieved robust quartile-based separation of overall and progression-free survival, with a PFS hazard ratio of 7.23 (highest vs. lowest quartile; p<0.0001). Prognostic performance was further confirmed with a previously reported fibrillar collagen model (COL210), identifying ECM remodeling as the biological driver of the peptide alterations depicted in urine in association with BC progression. These classifiers provide a reproducible, urine-based approach to non-invasive survival prediction, enabling risk-adapted surveillance and earlier therapeutic intervention. While urinary peptidomics requires prospective validation, it is poised to reduce reliance on invasive cystoscopy and to advance personalized care through clinical implementation.

## Introduction

Bladder cancer (BC) is one of the most common malignancies of the urinary tract and ranks among the top ten cancers worldwide, with over 570000 new cases and 200000 deaths annually (1). Although incidence rates are higher in men, women tend to present with more advanced disease and experience poorer outcomes (2). Well-established risk factors include tobacco smoking, occupational exposure to aromatic amines, chronic urinary tract inflammation, and certain genetic predispositions (3). More than 70% of newly diagnosed patients present with non-muscle-invasive BC (NMIBC). Initial treatment by transurethral resection is effective; however, recurrence occurs in up to 70% of cases within five years, and progression to muscle-invasive disease develops in 10–30% (4). These dynamics contribute to one of the highest lifetime disease burdens in oncology, significantly impacting patient morbidity and healthcare costs.

Cystoscopy remains the gold standard for diagnosis and surveillance, but it is invasive, costly, and limited in sensitivity, especially for flat or superficial lesions such as carcinoma in situ (5). The repeated nature of cystoscopies imposes a substantial physical, psychological, and financial burden on patients. Although urine cytology is specific for high-grade tumors, its sensitivity for low-grade disease is poor, and its overall performance in routine multicenter settings is lower than initially reported (6, 7). These methodological limitations underscore an important unmet need, the development of reliable, accurate, and non-invasive biomarkers that complement cystoscopy and improve patient-centered risk prediction.

To address recurrence and progression risk, clinicopathologic calculators like the European Organization for Research and Treatment of Cancer (EORTC) tables and the Spanish Club Urológico Español de Tratamiento Oncológico (CUETO) scoring model are widely used (8, 9). Although helpful in clinical decision-making, the EORTC tables often overestimate risk in BC patients managed according to current clinical practice, whereas CUETO tends to underestimate risk in those treated with intravesical Bacillus Calmette–Guérin (BCG) (10). Crucially, these models do not incorporate molecular heterogeneity, now regarded as a key driver of the diverse clinical course in BC. As a result, clinical tools under development incorporating molecular biomarkers are a promising avenue to refine prognostic accuracy and support personalized patient management.

Proteomic and peptidomic approaches have shown particular promise in biomarker discovery. Urine is an ideal biofluid for such studies. It is non-invasively collected, readily obtainable, stable at ambient conditions, and in direct contact with the urothelial lining (11). Over the past two decades, capillary electrophoresis coupled to mass spectrometry (CE–MS) has emerged as a robust, standardized platform for urinary biomarker analysis. CE–MS enables sensitive detection of hundreds of naturally occurring peptides in a single sample, offering a dynamic snapshot of physiological and pathological processes (12). Employing harmonized standard operating procedures, CE–MS has produced validated urinary biomarker panels such as BC-116 for primary detection and BC-106 for recurrence surveillance, with areas under the curve (AUC) of 0.82–0.87 in large multicenter studies, outperforming cytology and complementing cystoscopy (13, 14). These classifiers have been developed through reproducible pipelines, exploiting peptide deconvolution, normalization to standards, and machine-learning algorithms, with performance confirmed in independent cohorts and laboratories (13). Beyond diagnostic utility, urinary peptide signatures offer valuable biological insights. Multiple studies have consistently shown enrichment of peptides derived from collagens and other extracellular matrix (ECM) components in cancer-related urinary panels(15–17). This aligns with the fundamental role of ECM remodeling in tumor invasion, angiogenesis, stromal interactions, and therapeutic resistance (18). Aberrant protease activity, dysregulated collagen turnover, and fibrosis-like matrix accumulation contribute to progression from NMIBC to muscle-invasive disease (19). As ECM degradation products are filtered into urine, urinary peptidomics provides a direct, biologically relevant measure of tumor-stromal dynamics. Thus, collagen and ECM-derived urinary peptides are compelling candidates as both biomarkers and mechanistic readouts of disease biology.

Despite a strong biological rationale and the robust CE-MS methodology, up to now urine-based survival classifiers remained underexplored. Previous CE–MS research has focused mainly on diagnosis and recurrence detection, leaving a gap in prognostic applications (12, 20). Addressing this gap could improve risk stratification for overall survival (OS) and progression-free survival (PFS), and subsequently guide patients who may benefit from intensified adjuvant or systemic therapies, as well as enable reductions in surveillance for those at low risk, ultimately improving quality of life and reducing healthcare costs. In this study, we pursued a survival-focused urinary peptidomics strategy in BC by focusing on identifying and integrating survival-associated peptides into a machine learning classifier.

## Materials and Methods

### Study design and cohorts

A survival-oriented urinary peptidomics study was conducted in two independent BC cohorts, following the REMARK reporting recommendations and clinical proteomics guidelines(21, 22). Clinical Cohort-1 (n = 131) included previously acquired and reported urinary profiles aiming primarily at the detection of primary or recurrent BC (diagnostic setting) (12). This cohort was used for discovery and model development. Newly collected urine samples from an independent prospectively collected clinical cohort of patients with BC (Cohort-2; n = 102) were considered for external validation. Cohort-2 included patients attending “Laikon” General Hospital in Athens (Ethical approvals 565/25.16.2018 and 17554/ 22.11.2024). Eligible patients had: (i) a first bladder lesion detected on imaging, without previous pathological confirmation of urothelial BC, (ii) baseline urine available before any intervention, and (iii) follow-up information on OS and PFS events. Exclusion criteria were other active malignancies at baseline or inadequate urine volume/quality for CE–MS profiling. Urine was collected from 112 patients before cystoscopy or treatment according to institutional standard operating procedures (SOPs) and processed blinded to clinical outcomes. Out of 112, two were excluded failing the quality control (QC) criteria, and eight were excluded as there were missing follow-up data (**Supplementary Figure 1**). The complete workflow of the study is illustrated in **Figure 1**. To increase statistical power at discovery and stabilize peptide-level detection frequencies, we augmented the discovery group with peptidomics profiles from 96 subjects from a population study, retrieved from the Human Urinary Proteome Database (HUPD) (23). These profiles that served as control measurements were derived from anonymized subjects with no evidence of urological malignancy. All control samples were processed under identical CE–MS standard operating protocols and normalization, ensuring data comparability (20). Population controls were used exclusively for estimating peptide presence/absence frequencies, supporting multiple-testing control at the low-abundance end, and providing quality control anchors. They did not contribute to clinical events or survival time and were not included in the survival analyses. Basic demographics (age, sex, stage, and grade distribution) of cohorts and controls are summarized in **Supplementary Tables S1-S5.**

**Figure 1.**
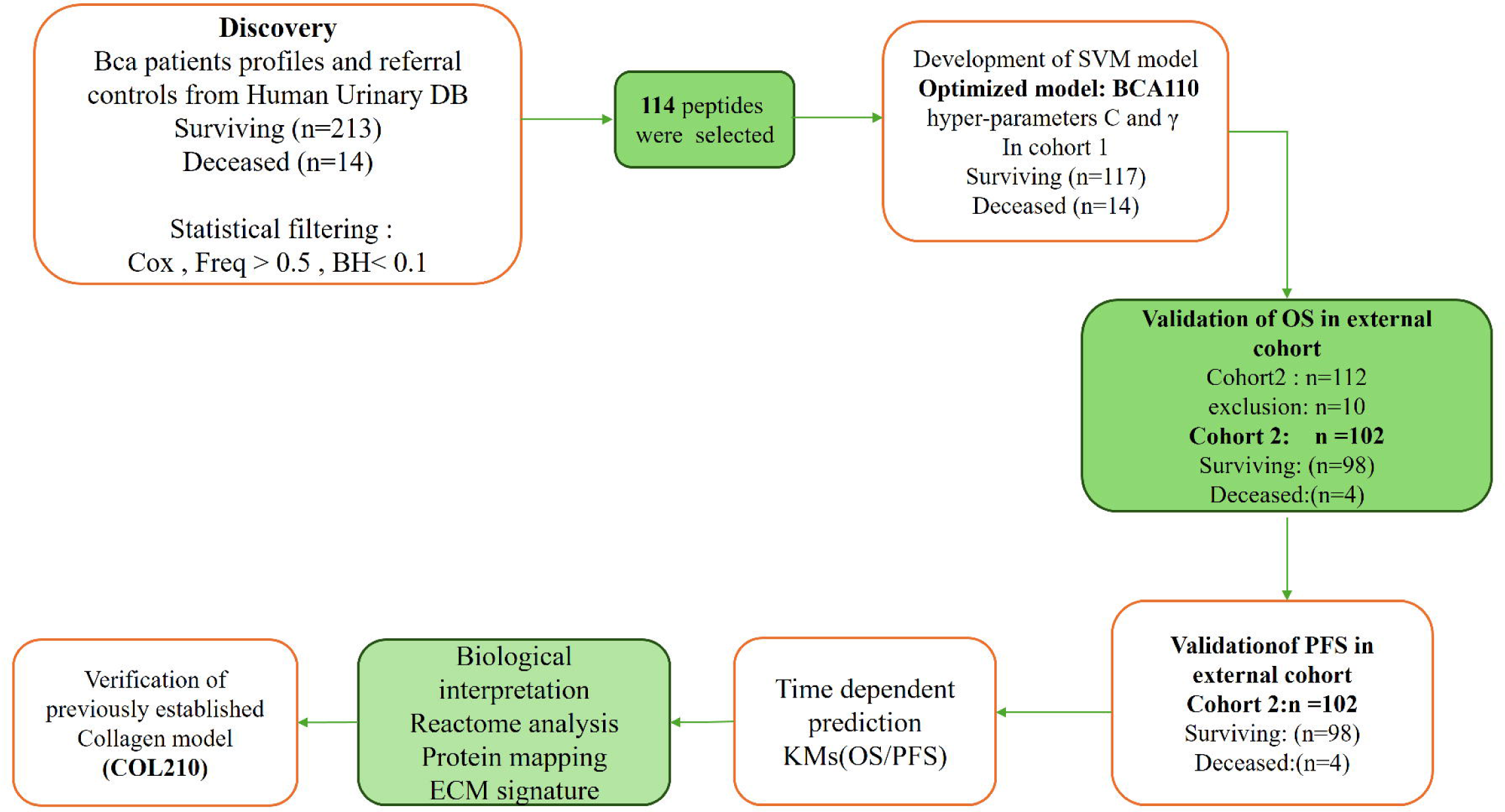
Workflow of the study for peptide biomarker discovery and model validation. Overview of the study design. Urinary peptide profiles from bladder cancer (Bca) patients were analyzed in the discovery cohort (surviving n = 213, deceased n = 14). Peptides were filtered based on Cox regression (p < 0.1, frequency > 0.5, Benjamini–Hochberg < 0.1), and 114 peptides were selected for model construction. An optimized SVM-based model (BC110) was developed and validated in independent cohorts for overall survival (OS) and progression-free survival (PFS). Biological interpretation, protein mapping, and comparison with the previously established COL210 collagen model were performed to assess ECM-related signatures.

### Urine collection _CE–MS sample preparation and acquisition

Voided midstream second urine of the morning was collected in sterile containers and stored at −80°C until further processing. Sample preparation followed established protocols (15, 24). Briefly, 700 µL of urine was diluted 1:1 with alkaline buffer (2 M urea, 10 mM NH□OH, 0.02% SDS; pH 10.5) and larger proteins were removed using a 20 kDa ultrafiltration device. The low molecular mass filtrate was desalted on PD-10 columns, lyophilized, and stored at 4 °C until analysis. Capillary electrophoresis (CE) was performed on a P/ACE MDQ system (Beckman Coulter) online-coupled to a microTOF-MS (Bruker Daltonics). Spectra were deconvoluted and molecular features defined by accurate mass, normalized migration time, and normalized intensity. Intensities were normalized to 29 endogenous collagen fragments as internal standards (19). Platform calibration, system suitability, and inter-batch monitoring were routinely implemented (25, 26).

### Peptide sequencing and database mapping

Representative samples were sequenced using CE–MS/MS with database searching against UniProt. Variable modifications included oxidation of methionine, hydroxylation of proline, and deamidation. False discovery rate (FDR) control was set at 1%, mass deviation was set to <5 ppm for MS and <50 mDa for fragment ions. Identified peptides were matched to curated urinary proteome resources, including the HUPD (23).

### Feature discovery and model development

In Cohort-1, peptides were first pre-filtered based on the missing signal levels, by requiring detection in at least 50% of samples. Survival association was then tested using univariable Cox proportional hazards models, and multiple-testing control was applied using the Benjamini– Hochberg false discovery rate (FDR) procedure. Candidate marker peptides significantly correlated with survival were combined to construct a composite classifier, through a support vector machine (SVM) framework (MosaCluster v1.7.0 (27), followed by in-house implementations in R/Python. Model optimization included a take-one-out cross-validation to assess the contribution of each peptide. In addition, a previously reported fibrillar collagen model (COL210) was investigated (28).

### Endpoints and statistical analysis

OS was defined as the time from urine collection to death. PFS was defined as time to first documented relapse/progression; deaths without relapse were censored. The index date was urine collection. Analyses were performed in R (v4.3.1) with the survival, survminer, and ROC packages. Exploratory analyses used Python (v3.11) with lifelines and matplotlib. Kaplan–Meier curves, log-rank tests, and Cox proportional hazards models were performed, with Schoenfeld residuals to test proportionality.

## Results

### Cohort characteristics

Two independent cohorts of patients with BC were analyzed. Cohort□1 (n=131) was used for discovery of significant peptide biomarkers and for subsequent modeling using machine learning, while Cohort□2 (n=102) for external validation. In Cohort-1, 14 OS events and 66 PFS events were observed, with a mean follow-up of 623 days. In Cohort-2, there were 4 OS events and 9 PFS events, with a mean follow-up of 1605 days **(Supplementary Table S6).**

### Discovery of survival□associated peptides and model development (BC110)

All urine samples were prepared and analysed using CE-MS. The datasets were calibrated against the standard set of internal reference peptides to ensure inter-sample comparability (20). In a first step, 213 control datasets (117 from BC patients without recurrence and 96 additional controls) were compared to 14 datasets from patients with recurrence to define peptides significantly associated with BC recurrence. This resulted in the identification of 114 potential biomarkers. Subsequently, the list of 114 CE-MS features (peptides) was then tested for association with survival using univariable Cox regression with resampling, while enforcing presence□frequency thresholds (≥50% of discovery samples or supported by population controls) and multiple□testing control (Benjamini–Hochberg, FDR < 0.10). Feature discovery was confined to the training set, yielding 114 urinary peptides significantly associated with OS. Full peptide□level statistics (p□values, detection frequencies) are reported in **Supplementary Table S7**. The 114 peptides were subsequently integrated using an SVM-based machine learning algorithm, and the model was further optimized. First, a grid search for SVM hyperparameters C and γ was performed based on complete take-one-out cross-validation. This resulted in an optimal C (640) and gamma (0.000008). In the next step, all peptides were investigated for their value in supporting classification; if classification was inferior in the presence of the respective peptide, this peptide was omitted. These steps resulted in an optimal classifier including 110 peptides (BC110). Model performance was first assessed using a complete take-one-out cross-validation of Cohort-1, yielding a ROC AUC of 0.80 (p value<0.001), indicative of good separation between survivors and non-survivors.

### Model performance and validation

External validation was conducted by applying the locked 110-peptide model directly to the test set without retraining. When the BC110 classifier was applied to the independent validation set (Cohort-2), the performance remained robust with an AUC of 0.78 (p value=0.03), supporting generalizability (**Supplementary Figure 2**).

### Functional enrichment reveals key collagen-related pathways

Functional enrichment analysis was performed to explore the biological context of the CE-MS– derived peptide panel. Out of 114 peptides, 42 high-confidence sequences mapping to 26 unique proteins were used for Gene Ontology (GO) and Reactome pathway enrichment **(Supplementary Table S8**). Over-representation analysis revealed that the majority of identified proteins were involved in extracellular matrix (ECM) organization and collagen-associated structural modules **(Figure 2A).** The top GO biological processes included collagen fibril organization, ECM structural constituent, and extracellular structure organization, with false discovery rates (FDRs) ranging from 1 × 10□¹□ to 1 × 10□□ **(Supplementary Table S9).** Reactome pathway enrichment further confirmed this pattern, highlighting collagen formation, collagen degradation, ECM proteoglycans, and integrin cell-surface interactions as the most significantly enriched pathways (all FDR < 1 × 10□□). Among these, collagen chain trimerization and assembly of collagen fibrils and other multimeric structures showed the strongest enrichment (FDR = 6.6 × 10□¹□), indicating a pronounced activation of collagen remodeling and stromal matrix turnover **(Table 1).** The protein–protein interaction (PPI) network constructed via STRING (**Figure 2B)** demonstrated a dense interaction hub dominated by COL1A1, COL1A2, COL3A1, COL5A2, COL12A1, and COL14A1, supporting the existence of a tightly connected collagen network. Peripheral nodes such as SRSF4, KHDRBS1, and HNRNPDL formed smaller sub-clusters related to RNA processing, suggesting potential post-transcriptional regulation within the same proteomic context. Consistent with ECM-driven tumor biology, the BC110 peptide set captures a tumor-associated ECM signature characterized by concurrent collagen biosynthesis, degradation, and structural reorganization hallmarks compatible with active tumor–stroma crosstalk in aggressive disease**. (Supplementary Table S10)**

**Figure 2A.**
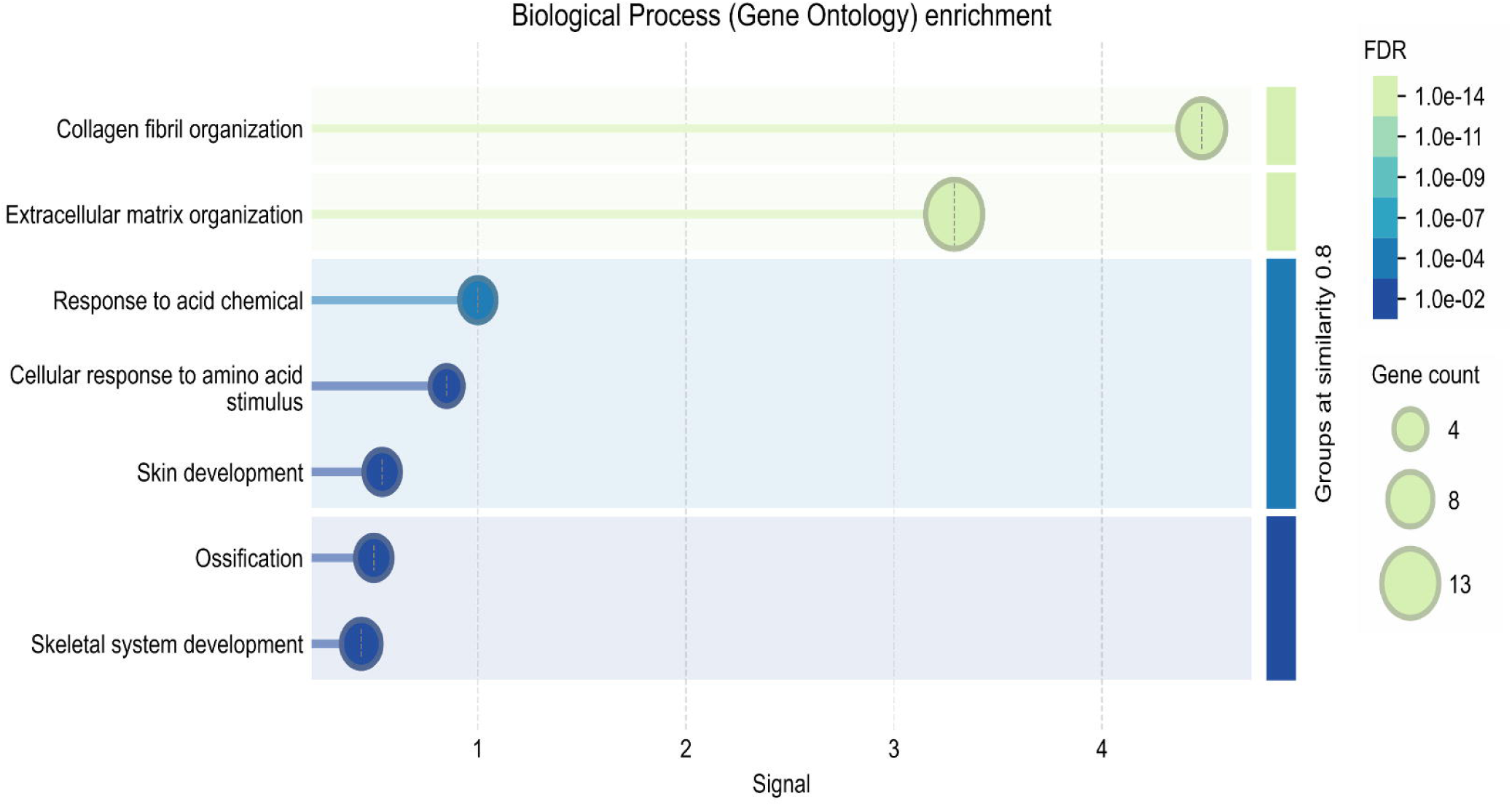
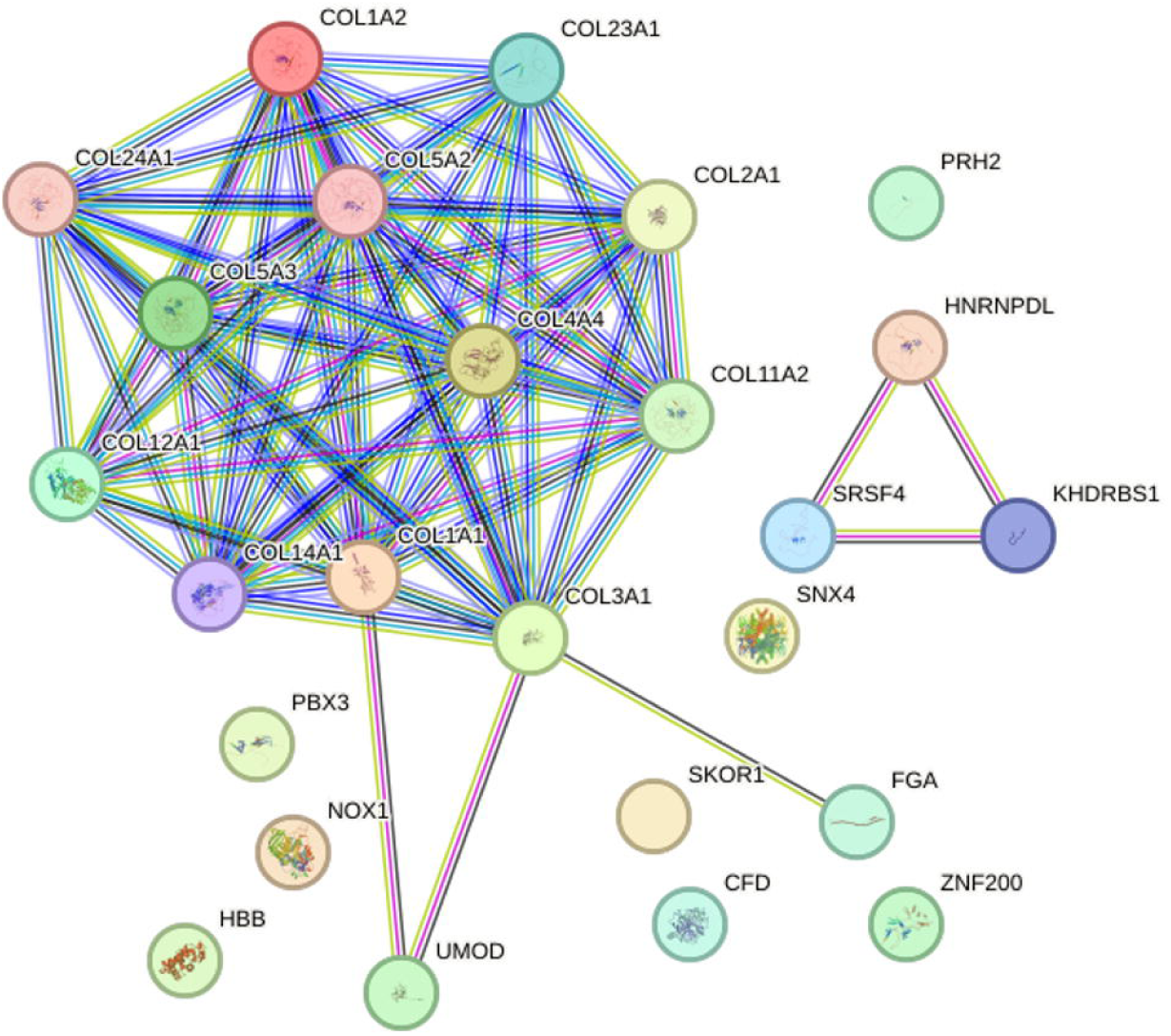
Gene Ontology (GO) enrichment analysis of proteins associated with the BC110 signature. Bubble plot showing significantly enriched biological processes based on proteins corresponding to the selected peptides. Collagen fibril and extracellular matrix organization were the most significantly enriched GO terms, followed by processes related to tissue development and chemical response. Bubble size represents the number of genes, and color intensity corresponds to the false discovery rate (FDR). **Figure 2B. Protein–protein interaction (PPI) network analysis using STRING.** Network visualization of proteins represented by the BC110 peptide panel. Highly interconnected collagen family members (COL1A1, COL1A2, COL3A1, COL5A2, COL14A1) form a central ECM-associated cluster, highlighting structural and signaling roles in extracellular matrix organization. Node size indicates connectivity degree; edge colors represent the type of evidence supporting interactions.

**Table 1.**
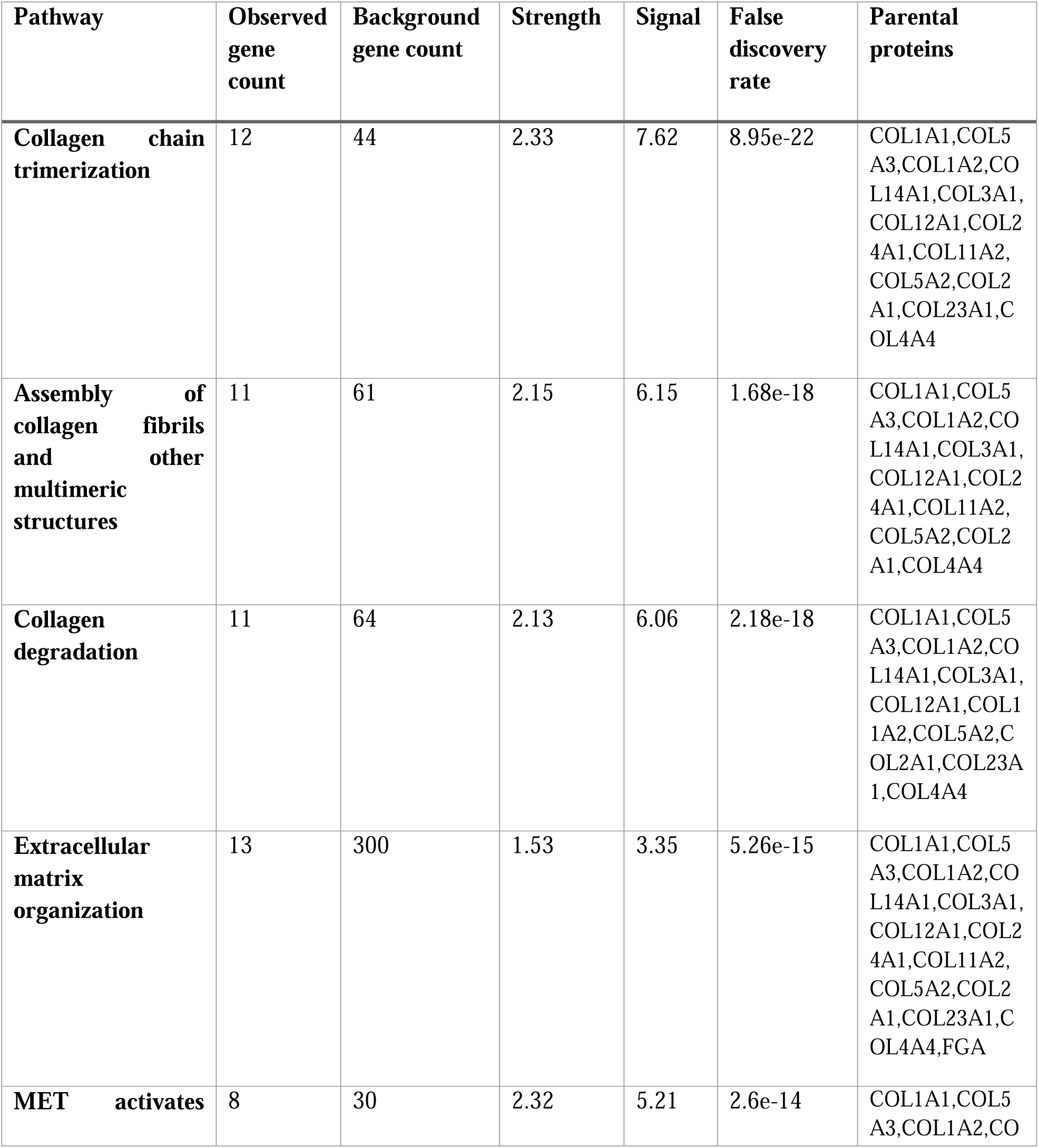
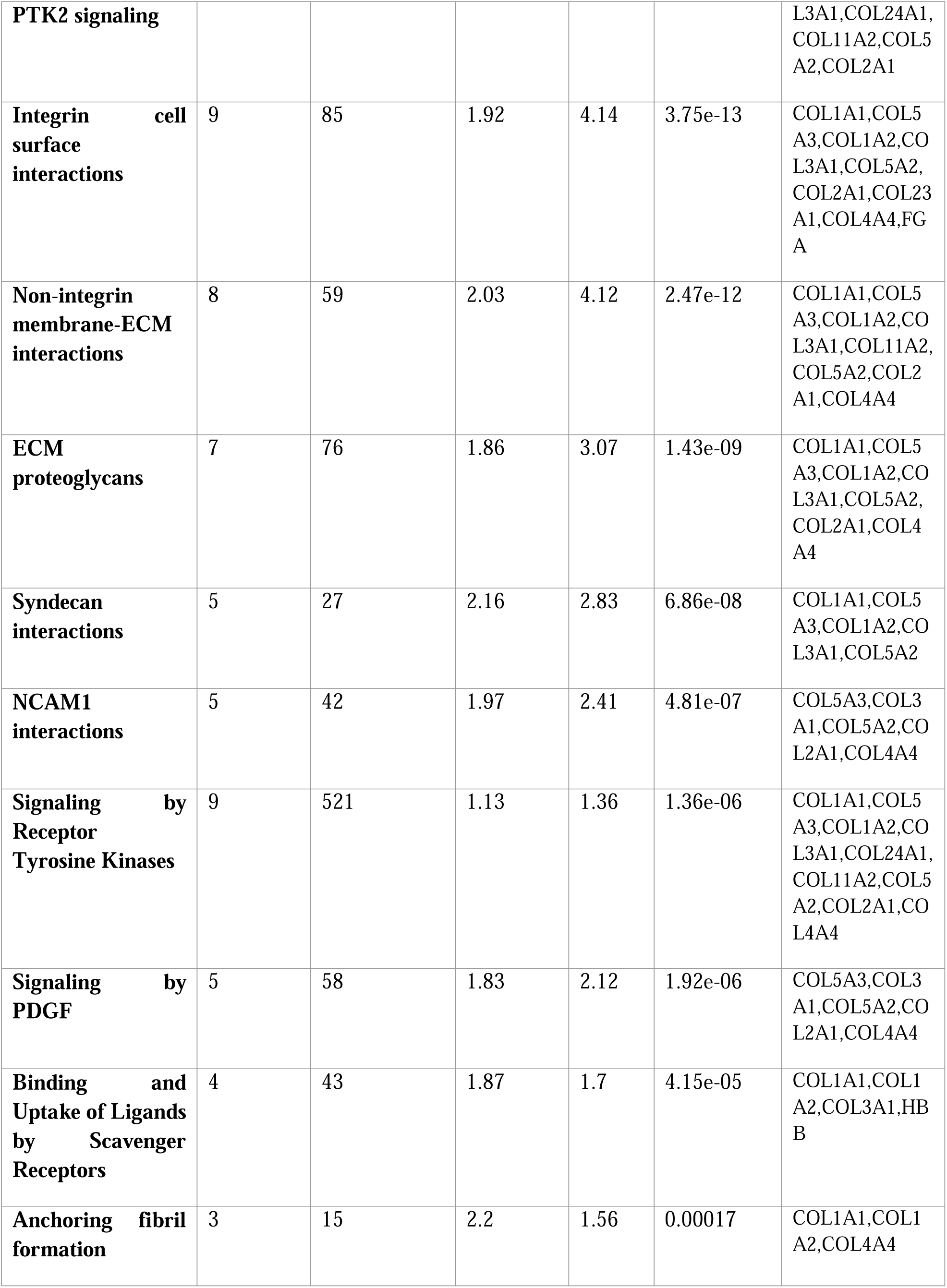
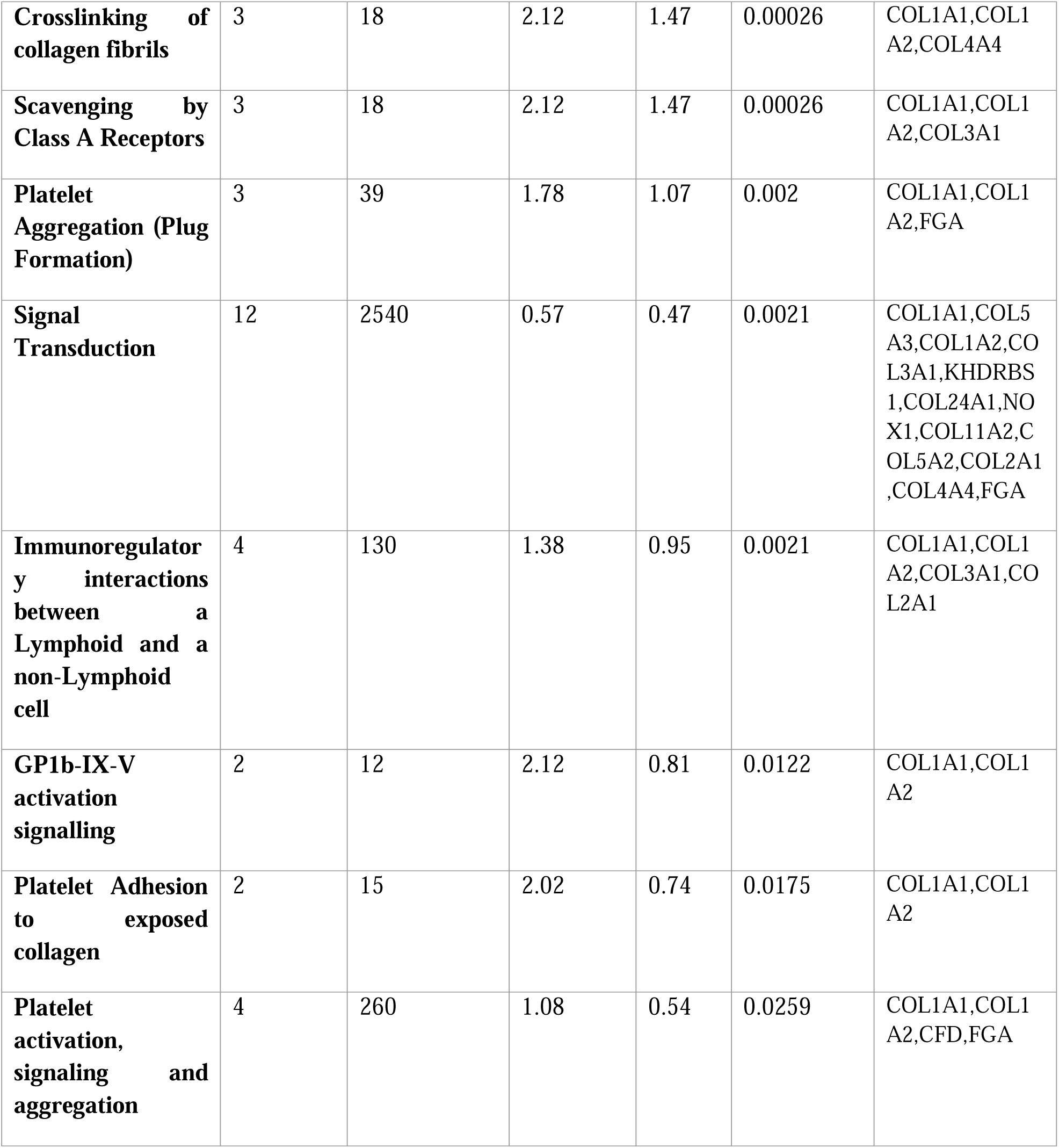
Reactome pathway enrichment.

### BC110 model predicts BC related outcomes

In a univariable Cox proportional hazards model including 233 patients from both independent cohorts (Cohort 1, n = 131; Cohort 2, n = 102), of whom 18 (7.73%) experienced an OS event, the BC110 score was significantly associated with overall survival (χ² = 33.21, df = 1, P < 0.0001). BC110 model demonstrated an excellent fit (−2 Log Likelihood: null model = 172.233, full model = 139.025). The regression coefficient for the BC110 score was b = 1.4345 ± 0.2248 (Wald χ² = 40.74, P < 0.0001), corresponding to a hazard ratio (HR) of 4.20 (95% CI: 2.70–6.52). BC110 model’s discriminative performance was high, with a Harrell’s C-index of 0.83 (95% CI: 0.74–0.93), indicating strong prognostic accuracy. Similarly, for progression-free survival (PFS), the BC110 score remained a significant predictor of outcome (χ² = 41.92, df = 1, P < 0.0001). The regression coefficient was b = 0.9930 ± 0.1435 (Wald χ² = 47.87, P < 0.0001), corresponding to a hazard ratio (HR) of 2.70 (95% CI: 2.04–3.58). The model showed significant discriminative performance with a Harrell’s C-index of 0.71 (95% CI: 0.66–0.77), indicating that higher BC110 scores were associated with shorter PFS. To further illustrate the prognostic stratification power of BC110, patients were grouped into quartiles (Q1–Q4) according to their BC110 classifier scores (n per quartile: 58, 59, 58, and 58). Kaplan–Meier analysis demonstrated a highly significant global separation of overall survival (log-rank χ² = 34.97, df = 3, P < 0.0001). Pairwise Cox proportional hazards comparisons indicated a stepwise increase in mortality risk, with Q1 vs Q3 HR = 6.38(95% CI 1.77–23.02) and Q1 vs Q4 HR = 20.54 (95% CI 5.06–83.31). No OS events were observed in Q2, precluding estimation relative to other quartiles (**Figure 3A).** When analyzed for PFS, stratification by the BC110 classifier similarly revealed marked and progressive differences in patient outcomes (log-rank χ² = 34.55, df = 3, P < 0.0001). Mean PFS times decreased gradually from Q1 to Q4 (1965, 1736, 1262, and 1037 days, respectively), reflecting a clear gradient in disease aggressiveness. Pairwise Cox contrasts confirmed increasing relapse risk in higher quartiles, with Q1 vs Q2 HR = 2.27 (95% CI 1.25–4.12), Q1 vs Q3 HR = 5.76 (95% CI 3.07–10.83), and Q1 vs Q4 HR = 7.23 (95% CI 3.74–13.98). (**Figure. 3B).**

**Figure 3A.**
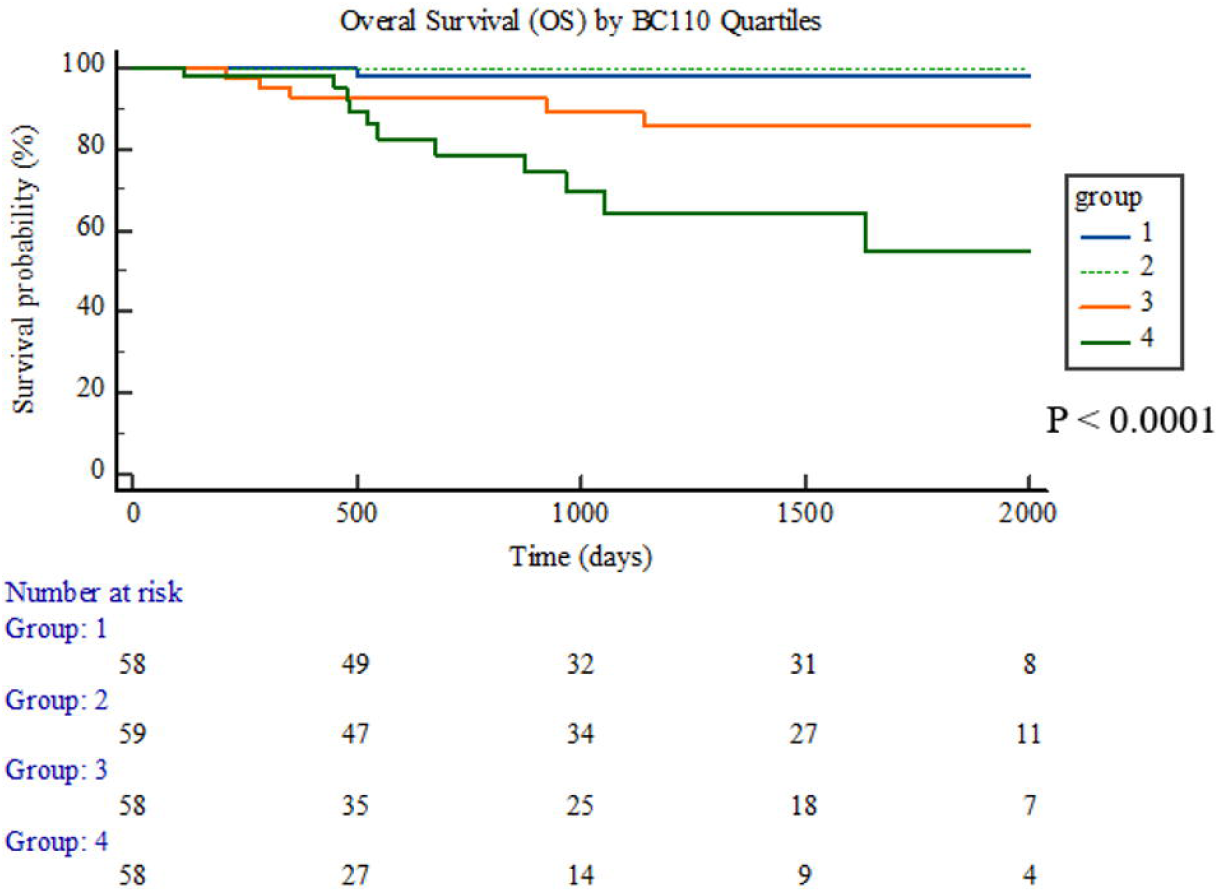
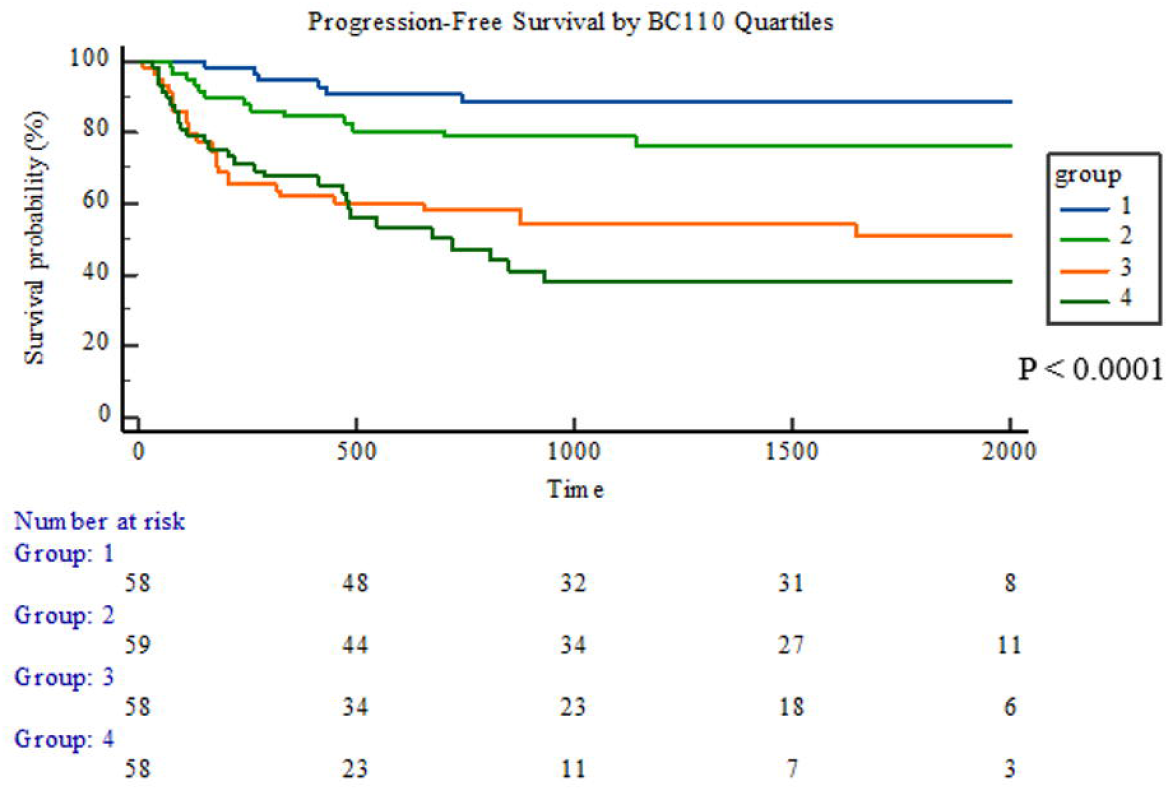
Kaplan–Meier analysis of overall survival (OS) by BC110 quartiles. Patients were stratified into four quartiles according to the BC110 SVM score. Higher BC110 scores were significantly associated with decreased overall survival (p < 0.0001, log-rank test). Numbers at risk for each group are indicated below the plot. **Figure 3B**. Kaplan–Meier analysis of progression-free survival (PFS) by BC110 quartiles. Patients were stratified into four quartiles according to the BC110 SVM score.Progression-free survival was analyzed. Patients with high BC110 scores exhibited significantly shorter PFS compared with lower-score groups (p < 0.0001). Numbers at risk for each quartile are displayed beneath the survival curves.

### Comparative prediction with the collagen-only peptide model

Given the collagen enrichment observed in the BC110 classifier, a previously reported collagen signature based solely on fibrillar collagens (COL210) was tested as an orthogonal comparator (28). In a univariable Cox proportional hazards model including 233 patients from both independent cohorts (Cohort 1, n = 131; Cohort 2, n = 102; 18 OS events), the COL210 score was significantly associated with overall survival (χ² = 22.67, df = 1, P < 0.0001; −2 Log Likelihood: null = 172.23, full = 149.57). The regression coefficient was b = 0.001817 ± 0.0003927 (Wald χ² = 21.41, P < 0.0001), corresponding to a hazard ratio (HR) = 1.01 per 1-unit increase (95% CI: 1.0010–1.0026). Discrimination was good (Harrell’s C-index = 0.80, 95% CI 0.70–0.90). A similar association was observed for progression-free survival (PFS), where the COL210 score remained a significant predictor (χ² = 19.39, df = 1, P < 0.0001; −2 Log Likelihood: null = 777.31, full = 757.92). The regression coefficient was b = 0.000848 ± 0.000190 (Wald χ² = 19.92, P < 0.0001), corresponding to a hazard ratio (HR) = 1.01 per 1-unit increase (95% CI: 1.0005–1.0012). Model discrimination for PFS was moderate (Harrell’s C-index = 0.64, 95% CI 0.58– 0.70). Similarly, when patients were stratified into quartiles according to their COL210 score (n per quartile: 59, 58, 58, and 58), Kaplan–Meier analysis again demonstrated significant global separation of OS (log-rank χ² = 15.34, df = 3, P = 0.0015). Pairwise Cox proportional hazards contrasts revealed stepwise increases in mortality risk with higher quartiles, with Q2 vs Q4 HR = 3.25 (95% CI 0.87–12.07) and Q3 vs Q4 HR = 2.16 (95% CI 0.51–9.22), while no deaths occurred in Q1(**Supplementary Figure 3**). For PFS, quartile stratification by the COL210 score also yielded significant global differences (log-rank χ² = 16.22, df = 3, P = 0.0010), showing a gradual decline in mean PFS from Q1 to Q4 (1869, 1575, 1335, and 1190 days, respectively). Stepwise increases in relapse risk were observed, with Q1 vs Q2 HR = 2.17 (95% CI 1.18–4.00), Q1 vs Q3 HR = 3.19 (95% CI 1.70–6.01), and Q1 vs Q4 HR = 4.01 (95% CI 2.12–7.57; **Supplementary Figure 4**). Collectively, these results confirm that both BC110 and COL210 capture prognostic information consistent with extracellular matrix–driven tumor biology, with BC110 demonstrating stronger discriminatory performance across both survival endpoints(**Supplementary Tables S11-S12).**

## Discussion

This study shifts the focus from detection to prognosis and shows that urinary peptide classifiers can stratify survival risk in BC using a non□invasive sample obtained before intervention. Here, we present a solid prediction of BC related outcomes based on a graded, quartile□wise classification that was derived by collagen-related peptides through machine learning. The prediction was most consistent for PFS in the combined cohort (n = 233; global log-rank p < 0.0001; Q1 vs Q4 HR = 7.23, 95 % CI 3.74–13.98)(28–30). Several urine-based and clinical prognosticators have been proposed in BC. Urinary tumour DNA (utDNA) approaches, particularly those tracking mutations as minimal residual disease (MRD) after cystectomy or using tumour-informed NGS panels, have demonstrated significant associations with disease progression and survival. Liu *et al* investigated 276 patients in a multicenter prospective study using tumour-informed next-generation sequencing (NGS) for urinary ctDNA detection after surgical treatment, and reported a hazard ratio (HR) of 8.3 for MRD-positive versus MRD-negative patients, with a concordance index (c-index) of 0.76 across both NMIBC and MIBC cohorts(31). Kim *et al* reported similar findings in a prospective cohort, confirming that utDNA positivity predicted recurrence and inferior disease-free survival independently of clinicopathologic variables(32). In a small pilot study, urinary IL-6 and IL-8 showed a significant increase with recurrence risk; however, these results need to be confirmed in a larger cohort(33). Cytogenetic assays such as UroVysion FISH have been incorporated into recurrence-risk nomograms, though discrimination remains moderate (C-indices around 0.70) (34). Traditional clinical tools such as the EORTC and CUETO scoring systems, while still widely used, show modest predictive accuracy in contemporary BCG-treated cohorts (C-indices of 0.60–0.66)(10). Our study provides a pre-intervention peptide-based classifier that integrates extracellular-matrix-derived collagen fragments into a machine-learning framework,comparable to the results obtained with utDNA. Our approach represents a biologically anchored urine-based prognostic tool that may complement existing utDNA and clinical risk models in predicting progression-free outcomes.

Biological annotation of the survival□associated peptides revealed a collagen□dominant extracellular matrix (ECM) signal, with a large fraction of the peptides mapping to COL1A1/1A2, COL3A1, and COL4A3/4. Notably, several central components in our panel have reported associations with bladder cancer, including COL1A1 and COL3A1 (see **Supplementary Table S10** for literature-based cancer links per gene). Gene Ontology and Reactome enrichment underscored ECM organization, collagen biosynthesis and degradation, and cell□matrix signaling modules. These observations align with the central role of ECM remodeling in tumor progression, invasion, and treatment resistance, wherein altered collagen architecture and protease activity (e.g., MMP□mediated cleavage cascades) facilitate migration and metastatic potential (35, 36). In urothelial carcinoma specifically, integrin–FAK signaling contributes to aggressive phenotypes and therapy resistance, providing a mechanistic bridge between matrix state and outcome (37, 38). These findings support the biological plausibility of our BC110 model in the context of bladder cancer.

A previously reported collagen□enriched comparator (COL210) using only fibrillar collagens, reproduced these trends (global log□rank p= 0.0010 for PFS; p= 0.0015 for OS). COL210 is an orthogonal, exclusively collagen□based model recapitulating survival separation, which strengthens the inference that the urinary peptidome is capturing microenvironmental remodeling rather than incidental analytical variation (28).

Our results extend a robust CE–MS literature that has primarily emphasized on diagnosis and recurrence detection (e.g., BC□116 and BC□106 panels) by demonstrating that survival□oriented peptide signatures can also be externally validated and clinically informative (12). The data suggest that the urinary peptidome indicates not only tumor presence but also associated biological processes, particularly high□turnover collagen dynamics relevant to prognosis. From a translational perspective, urine□based survival scores could complement guideline risk tools by flagging patients at high risk (upper quartiles) for intensified cystoscopic surveillance or earlier adjuvant/systemic evaluation, while enabling de□escalation for low□risk groups when appropriate. Decision□curve analysis and net□benefit frameworks provide standardized ways to quantify such trade□offs and should accompany prospective validation(39–41).

In contemporary BC care, progression/relapse events accrue earlier and more frequently than death, raising statistical power to detect gradients and yielding tighter confidence intervals. This is consistent with the behavior of time□to□event discrimination metrics under increasing follow□up (29, 30). Consistent with these reports, we observed a stronger discriminatory ability of the classifier for PFS versus OS in the study presented here, likely due to a larger number of endpoints and reduced distance in time to (urine) sampling of PFS in comparison to OS. The latter (OS events) were sparse, especially in Cohort□2, leading to wider pairwise CIs despite a significant global log□rank. Accordingly, PFS may be the pragmatic primary endpoint for initial clinical integration of the urine scores, with OS forming a confirmatory outcome as sample sizes and follow□up mature (42).

Evidence based on this study supports prediction of BC related outcomes in a group of patients primarily of non-muscle invasive, thus less advanced disease. Despite this promising evidence, this work presents some limitations. At first, event counts, particularly deaths, were modest, limiting precision for some contrasts. Moreover, although CE–MS workflows and internal□standard normalization mitigate analytical variability, our analysis is based on a single□platform; thereby cross□platform generalizability (e.g., LC–MS DIA) and inter□laboratory transfer cannot be assessed. However, an inter-laboratory comparison of the CE-MS-based CKD273 classifier reported in a previous study(20) gave very good results that also led to a letter-of-support from the US-FDA (https://www.fda.gov/media/99837/download). Additionally, the study lacks longitudinal urine sampling to test whether dynamic score changes track treatment response or molecular evolution and assess any potential environmental, treatment, or comorbidity influences on the urinary peptidome. Future work will proceed along three tracks, through a prospective, multi□center validation to confirm clinical utility (39, 40, 43). Second, mechanistic anchoring with tissue□level omics and imaging could link urine collagen fragments to tumor–stroma programs and matrix□modifying enzymes (e.g., LOX family) and quantify integrin/FAK pathway activation in matched specimens(35, 37, 44). Third, health□economic evaluations are needed to weigh assay costs against potential reductions in invasive cystoscopies and downstream resource use in NMIBC surveillance(45, 46). Together, these steps will help define where, when, and for whom urine□based survival scores meaningfully guide management in BC.

### Conclusions

Urinary CE–MS–based classifier (BC110) enables non-invasive risk stratification for both overall and progression-free survival in bladder cancer, capturing collagen-driven ECM remodeling biology. These signatures could complement guideline-based risk tools by refining surveillance intensity and identifying candidates for earlier systemic intervention. Prospective, multi-center validation and integration with established calculators are warranted before clinical implementation.

### Data Availability Statement

The data that support the findings of this study are openly available in Zenodo at https://doi.org/10.5281/zenodo.17397335.

## Supporting information

Supplementary Table S7, List of 114 peptides used for model construction (BC110)

Supplementary Table S9, GO enrichment analysis results

## Abbreviations

BC: Bladder cancer
CE-MS: Capillary electrophoresis-mass spectrometry
ECM: Extracellular matrix
OS: Overall survival
PFS: Progression-free survival
NMIBC: non-muscle-invasive BC
EORTC: European Organization for Research and Treatment of Cancer
CUETO: Club Urológico Español de Tratamiento Oncológico
BCG: Bacillus Calmette–Guérin
KM: Kaplan-Meier
HR: Hazard ratio
CI: Confidence interval
AUC(t): Time-dependent area under the ROC curve at time
SVM: Support vector machine
IQR: Interquartile range
QC: Quality control
FDR: False discovery rate
GO: Gene Ontology
HUPD: Human Urinary Proteome Database
IRB: Institutional Review Board
LC-MS/MS: Liquid chromatography-tandem mass spectrometry
REMARK: Reporting Recommendations for Tumor Marker Prognostic Studies
ROC: Receiver operating characteristic
SOP: Standard operating procedure
PDGF: Platelet-derived growth factor
FAK: Focal adhesion kinase

## Acknowledgments

All authors are grateful to all patients who donated urine samples. The authors would also like to thank Prof. Oleksii Rukhlenko, Dr. Sarah Robertson, and Prof. Boris N. Kholodenko for their valuable discussions, constructive feedback, and insightful comments that contributed to the development of this work. This work was supported by funding through the European Union’s Horizon Europe MULTIR (HORIZON-MISS-2023-CANCER-01-01, project number: 101136926) to MF, HM, and AV, funded by the European Commission. Views and opinions expressed are, however, those of the author(s) only and do not necessarily reflect those of the European Union. Neither the European Union nor the granting authorities HORIZON-MISS-2023-CANCER-01-01 can be held responsible for them. We also gratefully acknowledge the support of the Ministry of Science, Research and Technology of Iran (MSRT) and the Iran National Science Foundation (INSF).

## Conflict of interest statement

HM is the cofounder and co-owner of Mosaiques Diagnostics (Hannover, Germany), and MF is an employee of Mosaiques Diagnostics. All other authors declare no competing interests. AI tools were not used to generate hypotheses, analyze data, draw conclusions, and/or create figures. During the preparation of this manuscript, the authors employed generative AI and AI-assisted tools solely for language-level refinement.

## Supplementary Tables

**Supplementary Table S1.** Demographics and clinicopathologic summaries

**Supplementary Table S2.** Grade distribution of cohort 1

**Supplementary Table S3.** Stage and grade distribution of cohort 1

**Supplementary Table S4.** Tumor Stage Distribution of cohort 2

**Supplementary Table S5.** Tumor Grade Distribution in cohort 2

**Supplementary Table S6.** Cohort□level survival metrics

**Supplementary Table S7.** Full peptide□level statistics (p□values, detection frequencies) and detailed sequence and protein annotation

**Supplementary Table S8.** Protein list and mapping summary

**Supplementary Table S9**. Gene Ontology (GO) enrichment analysis

**Supplementary Table S10.** Cancer associations for the 26□protein panel

**Supplementary Table S11.**Cox models

**Supplementary Table S12.** Global log□rank statistics (quartiles)

## Notes

### Competing Interest Statement

HM is the cofounder and co-owner of Mosaiques Diagnostics GmbH (Hannover, Germany).
MF is employee of Mosaiques Diagnostics.
All other authors declare no competing interests.AI tools were not used to generate hypotheses, analyze data, draw conclusions, and/or create figures. During the preparation of this manuscript, the authors employed generative AI and AI-assisted tools solely for language-level refinement.

### Funding Statement

This work was supported in part by funding through the European Unions Horizon Europe MULTIR (HORIZON-MISS-2023-CANCER-01-01, project number: 101136926) to MF, HM, and AV, funded by the European Commission. Views and opinions expressed are, however, those of the author(s) only and do not necessarily reflect those of the European Union. Neither the European Union nor the granting authorities HORIZON-MISS-2023-CANCER-01-01 can be held responsible for them.

### Author Declarations

This study used human urine samples collected from bladder cancer patients, corresponded to a prospective study conducted at "Laikon" General Hospital in Athens, Greece, under the Ethics approval ID: 565/25.16.2018; 17554/ 22.11.2024. Ethical review and approval are not required for secondary use of previously reported datasets, due to all data being fully anonymized, based on the opinion of the ethics committee of the Hannover Medical School, Germany (no. 3116-2016).

