## Supplementary Information, Additional figures, and data tables for "Urinary peptidomic signatures predict overall and progression-free survival in patients with bladder cancer"

Table S1. Demographics and clinicopathologic summaries

| Cohort | N | Age, median years[IQR] | Male, n (%) | Female, n (%) |
| --- | --- | --- | --- | --- |
| Cohort 1 | 131 | 66.0 [58.0–74.0] | 102 (79.7%) | 26 (20.3%) |
| Cohort 2 | 102 | 69.0 [61.2–75.0] | 79 (77.5%) | 23 (22.5%) |
| HF controls | 96 | 61.0 [56.0–67.0] | 47 (56.6%) | 36 (43.4%) |

Table S2. Grade distribution of cohort 1

| Grade | N | % |
| --- | --- | --- |
| 2 | 28 | 37.84 |
| 3 | 18 | 24.32 |
| 1 | 12 | 16.22 |
| x | 2 | 2.7 |

Table S3.stage and grade distribution of cohort 1

| Stage | N | % |
| --- | --- | --- |
| pTaG2 | 23 | 31.08 |
| pTaG1 | 12 | 16.22 |
| pT0 | 10 | 13.51 |
| pT2G3 | 7 | 9.46 |
| pT1G3 | 7 | 9.46 |
| pT1G2 | 4 | 5.41 |
| pTaG3 | 3 | 4.05 |
| pTaGx | 2 | 2.7 |
| pT2G2 | 1 | 1.35 |
| pTxG3 | 1 | 1.35 |
| pTis | 1 | 1.35 |
| pTx | 1 | 1.35 |

Table S4:Tumor Stage Distribution of cohort 2

| Stage | N | % |
| --- | --- | --- |
| Ta | 51 | 50.0 |
| T1 | 41 | 40.2 |
| T2 | 9 | 8.8 |
| No tumor | 1 | 1.0 |

Table S5.Tumor Grade Distribution in cohort 2

| Grade Level | Type | N | % |
| --- | --- | --- | --- |
| Grade 2 | Low Grade | 51 | 50.0 |
| Grade 3 | High Grade | 21 | 20.6 |
| Grade 1 | Low Grade | 12 | 11.8 |
| Grade 2 | High Grade | 12 | 11.8 |
| Grade 4 | — | 2 | 2.0 |
| Grade 1 | — | 1 | 1.0 |
| Grade 2 | — | 1 | 1.0 |
| No tumor | — | 1 | 1.0 |
| Grade 5 | — | 1 | 1.0 |

Table S6. Cohort‑level survival metrics

| Cohort | OS events, n | PFS events(n) | mean follow-up(days) |
| --- | --- | --- | --- |
| Cohort-1 (n=131) | 14 | 66 | 623 |
| Cohort-2 (n=102) | 4 | 9 | 1605 |

Table S7. Full peptide‑level statistics (p‑values, detection frequencies) and detailed sequence and protein annotation

Table S8. Protein list and mapping summary

| No. | Protein name | Gene symbol | UniProt ID | Main biological role |
| --- | --- | --- | --- | --- |
| 1 | Collagen alpha-2(I) chain | COL1A2 | P08123 | Structural component of type I collagen fibrils, ECM integrity |
| 2 | Collagen alpha-1(I) chain | COL1A1 | P02452 | Major fibrillar collagen, structural support |
| 3 | Collagen alpha-4(IV) chain | COL4A4 | Q9NQC8 | Basement membrane component |
| 4 | Collagen alpha-1(III) chain | COL3A1 | P02461 | Fibrillar collagen, tissue elasticity and repair |
| 5 | Collagen alpha-3(V) chain | COL5A3 | Q5KU26 | Regulation of collagen fibril formation |
| 6 | Collagen alpha-1(XII) chain | COL12A1 | Q99715 | ECM organization, fibril-associated collagen |
| 7 | Collagen alpha-1(XXIII) chain | COL23A1 | Q96P44 | Cell adhesion and ECM remodeling |
| 8 | Serine/arginine-rich splicing factor 4 | SRSF4 | Q08170 | RNA splicing regulation |
| 9 | KH domain-containing, RNA-binding, signal transduction-associated protein 1 | KHDRBS1 | Q07666 | RNA binding and post-transcriptional regulation |
| 10 | Collagen alpha-1(XIV) chain | COL14A1 | Q05707 | Fibril assembly, ECM stability |
| 11 | Collagen alpha-2(V) chain | COL5A2 | P05997 | Fibrillar collagen component |
| 12 | Collagen alpha-1(XXIV) chain | COL24A1 | Q8NFW1 | Fibril assembly, tissue development |
| 13 | Heterogeneous nuclear ribonucleoprotein D-like | HNRNPDL | Q14103 | mRNA processing and transport |
| 14 | NADPH oxidase 1 | NOX1 | Q9Y5S8 | Reactive oxygen species generation |
| 15 | SKI family transcriptional corepressor 1 | SKOR1 | Q6ZUJ8 | Transcriptional repression and signaling regulation |
| 16 | Sorting nexin-4 | SNX4 | O95219 | Endosomal membrane trafficking |
| 17 | Collagen alpha-1(II) chain | COL2A1 | P02458 | Cartilage structural collagen |
| 18 | Microtubule-associated serine/threonine-protein kinase 2 | MAST2 | Q6P0Q8 | Cytoskeletal signaling and stability |
| 19 | Pre-B-cell leukemia transcription factor 3 | PBX3 | P40426 | Transcriptional regulation, development |
| 20 | Hemoglobin subunit beta | HBB | P68871 | Oxygen transport |
| 21 | Collagen alpha-2(XI) chain | COL11A2 | P13942 | Cartilage ECM component |
| 22 | Uromodulin | UMOD | P07911 | Kidney tubular function and immune regulation |
| 23 | Zinc finger protein 200 | ZNF200 | Q9UDV6 | DNA binding and transcriptional control |
| 24 | Salivary acidic proline-rich phosphoprotein 1/2 | PRH2 | P02810 | Salivary gland secretion and mineral homeostasis |
| 25 | Fibrinogen alpha chain | FGA | P02671 | Blood coagulation and clot formation |
| 26 | Complement factor D | CFD | P00746 | Complement cascade activation |

Table S9. Gene Ontology (GO) enrichment analysis

Table S10.Cancer associations for the 26‑protein panel

| Protein | Gene symbol | Cancer type(s) | Evidence | Key reference (PMID/DOI) | Link |
| --- | --- | --- | --- | --- | --- |
| Collagen alpha-1(I) chain | COL1A1 | Colorectal cancer; Bladder cancer | Promotes metastasis via WNT/PCP signaling in CRC; Type I collagen associated with bladder cancer progression. | PMID:29393423; PMID:27655672 | https://pubmed.ncbi.nlm.nih.gov/29393423/; https://pubmed.ncbi.nlm.nih.gov/27655672/ |
| Collagen alpha-2(I) chain | COL1A2 | Gastric cancer (collagen family context); Bladder cancer (type I collagen) | Type I collagen network implicated in tumor invasion; broader collagen alterations linked to progression. | PMID:27655672; PMC8815231 | https://pubmed.ncbi.nlm.nih.gov/27655672/; https://pmc.ncbi.nlm.nih.gov/articles/PMC8815231/ |
| Collagen alpha-1(III) chain | COL3A1 | Bladder cancer | Overexpression correlates with poor prognosis and advanced stage. | DOI:10.18632/oncotarget.19733 | https://pmc.ncbi.nlm.nih.gov/articles/PMC5642573/ |
| Collagen alpha-2(V) chain | COL5A2 | Gastric cancer; Lung adenocarcinoma | Drives EMT and malignant phenotypes in GC; promotes ER stress and immune modulation in LUAD. | DOI:10.1515/med-2022-0593; PMC12146537 | https://pmc.ncbi.nlm.nih.gov/articles/PMC9843231/; https://pmc.ncbi.nlm.nih.gov/articles/PMC12146537/ |
| Collagen alpha-1(XII) chain | COL12A1 | Pancreatic cancer; Intrahepatic cholangiocarcinoma; Breast cancer | High expression predicts poor prognosis and is CAF-enriched in pancreatic cancer; Upregulated and prognostic in iCCA; Linked to poor outcomes in breast cancer. | PMID:36900272; DOI:10.1186/s13148-022-01413-5; DOI:10.1530/ERC-23-0012 | https://pubmed.ncbi.nlm.nih.gov/36900272/; https://clinicalepigeneticsjournal.biomedcentral.com/articles/10.1186/s13148-022-01413-5; https://erc.bioscientifica.com/view/journals/erc/30/5/ERC-23-0012.xml |
| Collagen alpha-1(XIV) chain | COL14A1 | Breast cancer (premetastatic niche); Liver cancer | ECM remodeling with increased fibrillar collagens including COL14A1 primes metastasis; COL14A1 promotes self‑renewal in liver cancer models. | DOI:10.1038/s41598-023-45832-7; DOI:10.1097/JBR.0000000000000087 | https://www.nature.com/articles/s41598-023-45832-7; https://spj.science.org/doi/10.1097/JBR.0000000000000087 |
| Collagen alpha-1(XXIII) chain | COL23A1 | Clear cell renal cell carcinoma; Thyroid cancer (bioinformatics) | Oncogenic role promoting adhesion/invasion in ccRCC; dysregulated in thyroid cancer cohorts. | PMC5575106; ResearchGate (2024) | https://pmc.ncbi.nlm.nih.gov/articles/PMC5575106/; https://www.researchgate.net/publication/382953490 |
| Collagen alpha-1(XXIV) chain | COL24A1 | Gastric cancer (collagen-high subtypes) | Collagen-based subtyping of GC implicates multiple collagen genes including COL24A1 in tumor biology. | PMC11212290 | https://pmc.ncbi.nlm.nih.gov/articles/PMC11212290/ |
| Collagen alpha-4(IV) chain | COL4A4 | Breast cancer (ECM/basement membrane context) | Basement membrane collagens participate in tumor invasion and microenvironment remodeling. | Review (2023) PMC10007858 | https://pmc.ncbi.nlm.nih.gov/articles/PMC10007858/ |
| Collagen alpha-1(II) chain | COL2A1 | Pan-cancer ECM remodeling (context) | Fibrillar collagen implicated in tumor stiffness and invasion across solid tumors. | PMC10159947 | https://pmc.ncbi.nlm.nih.gov/articles/PMC10159947/ |
| Collagen alpha-2(XI) chain | COL11A2 | Gastric cancer (COL4A family/ECM) | Collagen family members show prognostic/therapeutic relevance in gastric cancer datasets. | DOI:10.21037/tcr-20-517 | https://tcr.amegroups.org/article/view/42315/html |
| Serine/arginine-rich splicing factor 4 | SRSF4 | Pan‑cancer (splicing factor dysregulation) | Splicing factors (including SRSF family) are recurrently altered and contribute to tumor progression and therapy response. | PMC10049280 (review on hnRNPs/splicing) | https://pmc.ncbi.nlm.nih.gov/articles/PMC10049280/ |
| KH domain-containing, RNA-binding, signal transduction-associated protein 1 | KHDRBS1 (Sam68) | Colon; Prostate; HCC; Pan‑cancer | Sam68 promotes proliferation/metastasis; high expression linked to poor prognosis; functional oncogenic roles in CRC and HCC. | eLife 2016; PMID:39450529; PMC11537268 | https://elifesciences.org/articles/15018; https://pubmed.ncbi.nlm.nih.gov/39450529/; https://pmc.ncbi.nlm.nih.gov/articles/PMC11537268/ |
| Heterogeneous nuclear ribonucleoprotein D-like | HNRNPDL | Cervical; Colorectal; Leukemia (CML) | Aberrant expression reported in CRC and cervical cancer; modulates growth and drug response via PBX1 in CML models. | NCBI Gene (2025 summary); DOI:10.1016/j.plantsci.2018.12.031 | https://www.ncbi.nlm.nih.gov/gene/9987; https://www.sciencedirect.com/science/article/abs/pii/S0378111918311715 |
| NADPH oxidase 1 | NOX1 | Colorectal cancer | Overexpression linked to oxidative stress, proliferation and tumor growth; silencing reduces CRC growth. | PMCID:PMC12174529; DOI:10.1016/j.yonc.2023.08.007 (example) | https://pmc.ncbi.nlm.nih.gov/articles/PMC12174529/; https://www.sciencedirect.com/science/article/pii/S2213231723002288 |
| SKI family transcriptional corepressor 1 | SKOR1 | Breast cancer (limited evidence) | SKOR1 implicated in TGF-β/SMAD regulation; reports of altered expression in some tumors (exploratory). | — |  |
| Sorting nexin-4 | SNX4 | Clear cell renal cell carcinoma | SNX4 expression correlates with immune infiltration and prognosis in ccRCC. | PMCID:PMC11424112 | https://pmc.ncbi.nlm.nih.gov/articles/PMC11424112/ |
| Microtubule-associated serine/threonine-protein kinase 2 | MAST2 | Liver cancer; Breast cancer (rearrangements) | MAST2 overexpression supports tumor growth; rearrangements reported in invasive breast carcinoma. | PMCID:PMC10419289; NCBI Gene (MAST2) | https://pmc.ncbi.nlm.nih.gov/articles/PMC10419289/; https://www.ncbi.nlm.nih.gov/gene/23139 |
| Pre-B-cell leukemia transcription factor 3 | PBX3 | Acute myeloid leukemia; Cervical cancer; Glioma | High PBX3 predicts poor prognosis and drives malignant phenotypes across cancers. | Cancers (Basel) 2020; PMCID:PMC10849273 | https://www.mdpi.com/2072-6694/12/2/431; https://pmc.ncbi.nlm.nih.gov/articles/PMC10849273/ |
| Hemoglobin subunit beta | HBB | Colorectal liver mets; Multiple epithelial cancers | HBB expression supports ROS resistance; HBB-positivity linked to worse survival. | Nat Commun 2017; PMID:37239002 | https://www.nature.com/articles/ncomms14344; https://pubmed.ncbi.nlm.nih.gov/37239002/ |
| Uromodulin | UMOD | Renal cell carcinoma | UMOD genotype associated with more aggressive RCC clinicopathologic features. | PMID:28753889 | https://pubmed.ncbi.nlm.nih.gov/28753889/ |
| Zinc finger protein 200 | ZNF200 | Pan‑cancer (KRAB‑ZNF family context) | ZNF family members show roles in tumor progression; ZNF200 reported interacting with PRMT3 and harboring mutations in cancers; HPA shows cancer‑tissue expression. | PMC6442004 (KRAB‑ZNFs); Biochem J 2024; HPA | https://pmc.ncbi.nlm.nih.gov/articles/PMC6442004/; https://portlandpress.com/biochemj/article/481/23/1723/235232; https://www.proteinatlas.org/ENSG00000010539-ZNF200/cancer |
| Salivary acidic proline-rich phosphoprotein 1/2 | PRH2 | Oral disease context (diagnostic/biomarker); limited direct cancer evidence | PRH2 polymorphisms associate with oral health phenotypes; salivary PRPs explored for oral disease/cancer monitoring. | PMID:29191562; 2024 review on salivary diagnostics | https://pubmed.ncbi.nlm.nih.gov/29191562/; https://www.sciencedirect.com/science/article/pii/S2772906024001468 |
| Fibrinogen alpha chain | FGA | Pan‑cancer (coagulation–cancer axis) | FGA participates in hypercoagulability and tumor progression; elevated fibrinogen often correlates with poor outcomes (literature broadly supports). | — |  |
| Complement factor D | CFD (Adipsin) | Breast cancer; AML | Adipsin/CFD promotes tumor invasion via adipocyte–cancer interactions in breast cancer; CFD shows prognostic relevance in AML datasets. | DOI:10.1038/s41598-024-69476-3; PMCID:PMC11414840 | https://www.nature.com/articles/s41598-024-69476-3; https://pmc.ncbi.nlm.nih.gov/articles/PMC11414840/ |

Table S11. Cox models

| Model | Endpoint | HR (95% CI) | p-value |
| --- | --- | --- | --- |
| BC110 | OS | 4.20 (2.70–6.52) | <0.0001 |
| BC110 | PFS | 2.70 (2.04–3.58) | <0.0001 |
| COL210 | OS | 1.0018 (1.0010 – 1.0026) | <0.0001 |
| COL210 | PFS | 1.0008 (1.0005–1.0012) | <0.0001 |

Table S12. Global log‑rank statistics (merged cohorts/based on quartiles)

| Model | Endpoint | Chi-square | DF | p-value |
| --- | --- | --- | --- | --- |
| BC110 | OS | 34.97 | 3 | < 0.0001 |
| BC110 | PFS | 34.55 | 3 | < 0.0001 |
| COL210 | OS | 15.34 | 3 | = 0.0015 |
| COL210 | PFS | 16.22 | 3 | = 0.0010 |

Fig S1. Cohort 2_Exclusion criteria

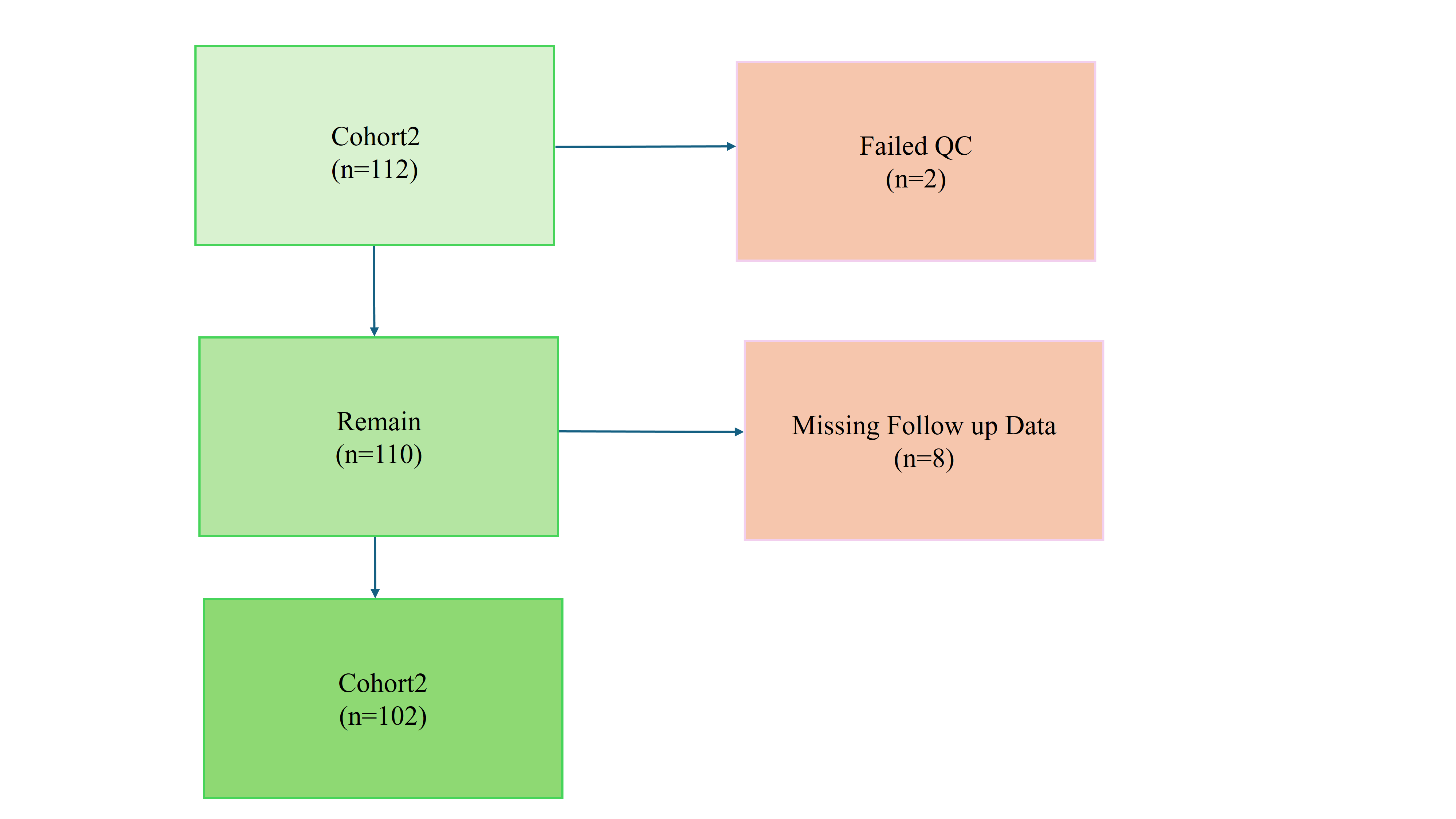

Fig S2. ROC curves (OS) for BC110 in cohort 2

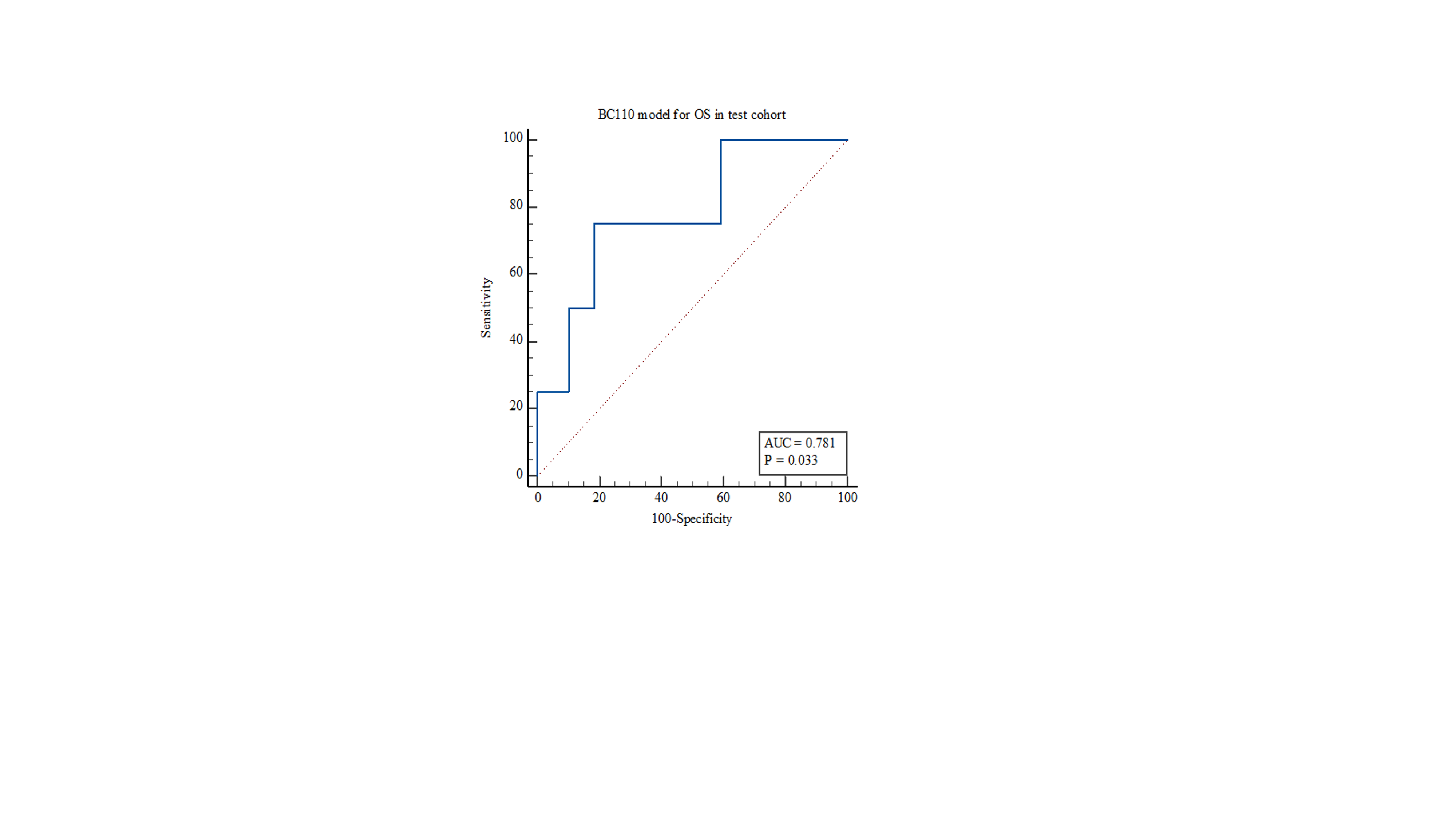

Figure S3. COL210 _Overall survival (quartiles)

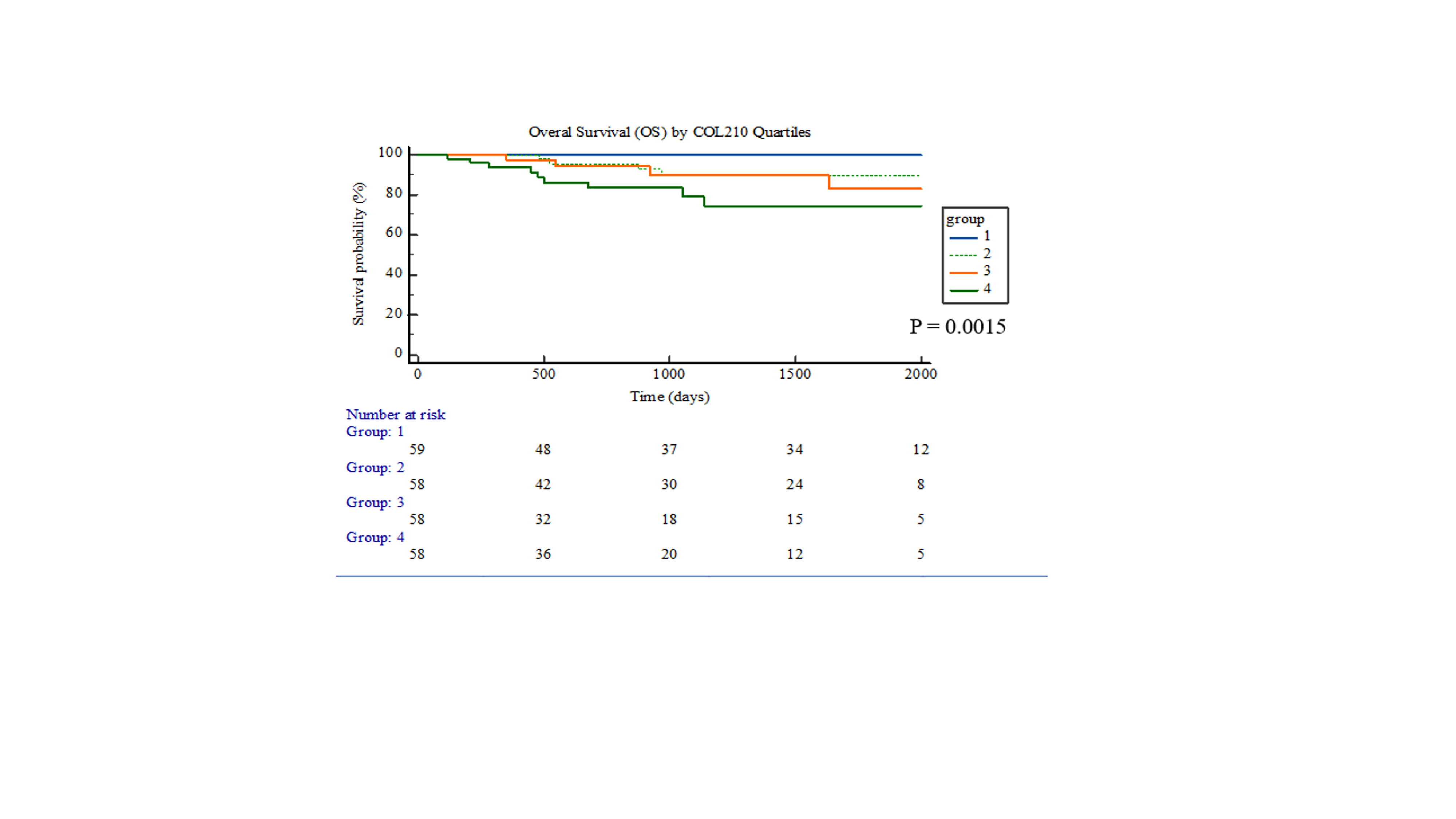

Figure S4. COL210 _Progression‑free survival (quartiles)

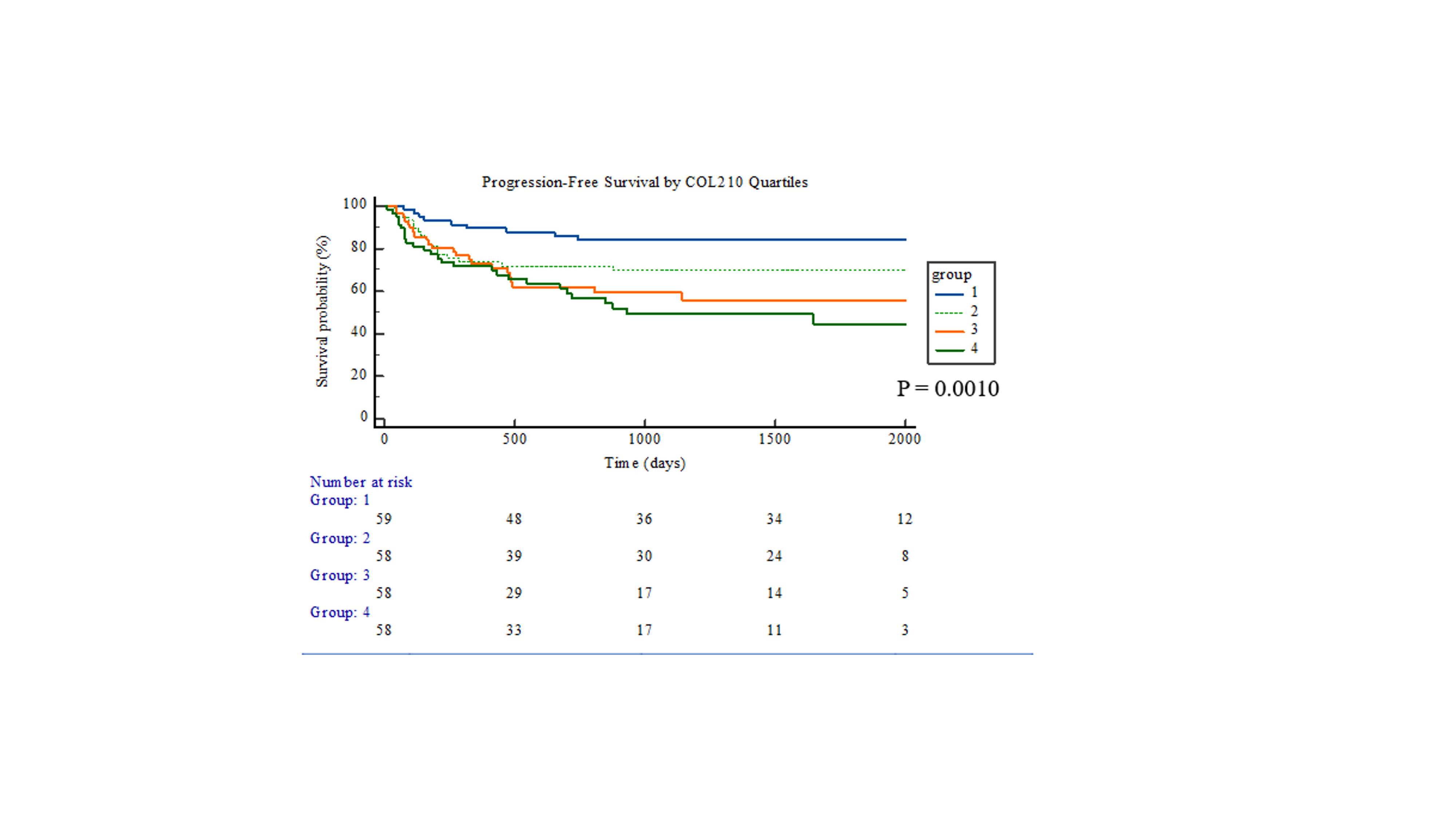
